# Histopathology-based Spatial Profiling of Immune and Molecular Features Predicts Cancer risk in Barrett’s Esophagus

**DOI:** 10.1101/2025.11.11.25339952

**Authors:** Caner Ercan, Xiaoxi Pan, Thomas G. Paulson, Matthew D. Stachler, Fahire Göknur Akarca, William M. Grady, Carlo C. Maley, Yinyin Yuan

## Abstract

**Background:** Improved cancer risk stratification is needed to differentiate high-risk individuals with Barrett’s esophagus (BE) from low-risk populations to reduce overtreatment and improve outcome. The evolution of BE towards adenocarcinoma is likely driven by a combination of genomic and microenvironmental factors, yet existing predictive models rarely integrate both using routine specimens.

**Method:** We developed BEACON (Barrett Esophagus DNA content Abnormality and immune ecology for Cancer Outcome), a spatially aware framework predicting DNA content abnormalities and characterizing immune spatial ecology from routine histopathology. First, using 777 BE biopsies with flow cytometry-based DNA content data scanned at two institutions, we trained and tested DACOR (DNA content abnormality recognition), a multi-instance learning model that predicts DNA content abnormalities from histopathology. Next, complementary models for cell classification and tissue segmentation enabled spatial immune ecology metric computations. Lastly, a logistic regression model integrated molecular immune ecological features and epithelial morphology for cancer risk stratification.

**Results:** DACOR achieved 0.825 AUC in the test cohort for DNA content abnormality prediction. DNA content abnormal regions exhibited increased lymphoplasma cellular inflammation versus normal regions (p=0.006). Patients classified as DNA content abnormal by DACOR demonstrated increased cancer progression (p=0.0001). Among patients with DNA content abnormality, cancer progressors exhibited increased plasma cell clustering adjacent to abnormal epithelium compared to non-progressors. The integrated risk classification model stratified DNA content abnormal patients into high- and low-risk groups with 0.817 AUC.

**Conclusion:** BEACON spatially integrates molecular abnormality with immune spatial ecology to stratify BE patients by cancer progression risk using routine pathology images. This scalable, explainable approach could improve clinical decision-making and reduce unnecessary surveillance in low-risk patients.

## Introduction

Surveillance of Barrett’s esophagus (BE) represents an important burden on healthcare systems,^1^ with increasing prevalence in Western populations.^2^ While monitoring precancerous lesions is important, the actual progression rate to esophageal adenocarcinoma (EAC) remains low.^3^ The growing BE population requiring surveillance far exceeds those who will progress, creating an urgent need for improved risk stratification to identify high-risk patients requiring intensive monitoring while reducing unnecessary surveillance in low-risk individuals.

DNA content abnormalities have been implicated in the development of EAC in BE^4,5^, and may be the most predictive single molecular biomarker for cancer progression risk.^6,7^ However, BE is molecularly heterogeneous, with DNA content abnormalities typically detected in only a minority of cells and exhibiting spatial heterogeneity.^8^ Beyond simply the presence, the spatial distribution patterns of these alterations can provide prognostic information in BE.^9^ Current methods for spatially characterizing DNA content are limited by labor intensive workflows, and the need for specialized equipment, leading substantial barriers to scalability, clinical implementation. This underscores the urgent need for spatially aware DNA content detection methods that can be applied to routine clinical specimens.

Progression to carcinoma is a dynamic evolutionary process in which genomic alterations in epithelial cells and adaptive immune responses interact continuously.^10^ While biopsies offer only static snapshots, they capture essential hallmarks of epithelial-immune co-evolution. Spatial molecular heterogeneity, immune cell diversity, and tissue organization can be interrogated using ecological analysis, which assesses architecture and cellular interactions to provide prognostic insights.^11^

Deep learning (DL) in computational pathology enables detection of molecular alterations directly from routine histopathology images. While DL studies in BE have focused on tissue classification,^12^ recent advances in other cancers demonstrate prediction of molecular features from H&E-stained slides while capturing spatial heterogeneity.^13^ Multi-instance learning (MIL) frameworks generate slide-level predictions while preserving spatial context, enabling identification of heterogeneous features such as regional aneuploidy.^14^

Beyond molecular alterations, the immune microenvironment plays a crucial role in shaping cancer progression.^15,16^ Computational pathology enables comprehensive spatial immune analysis, from density quantifications to sophisticated ecological metrics capturing diversity, organization, and cellular interactions.^11,17,18^ These immune ecological features demonstrate prognostic value in precancerous lesions,^18–20^ suggesting that the spatial architecture of immune responses may reflect or modulate progression risk.

In BE, the interplay between molecular alterations such as aneuploidy and the immune microenvironment remains incompletely understood. Evaluating both features spatially could provide complementary information for risk stratification. In this study, we present BEACON (Barrett Esophagus DNA content and immune ecology for Cancer Outcome), a computational pathology framework that integrates DL-based prediction of DNA content abnormality with spatial analysis of the immune ecology. We first train DACOR (DNA content abnormality recognition), using MIL to detect DNA content abnormalities from routine H&E-stained biopsy images. We then extract immune ecological metrics to characterize the immune landscape and its association with progression risk, and combine predicted DNA content status with epithelial morphology and immune features to stratify high-risk patients. By leveraging routine slides and avoiding tissue-destructive assays, this approach offers a practical, scalable strategy for cancer risk assessment in BE that captures both molecular heterogeneity and immune landscape features.

## Methods

### Dataset

H&E-stained whole slide images (WSIs) of endoscopic biopsies sourced from a subset of 191 BE patients from the Seattle Barrett’s Esophagus Annotated Resource (BEAR) collected following the Seattle protocol were utilized. (Figure 1A) Fresh tissue biopsies were divided into halves: one underwent flow cytometric DNA content assessment,^4^ while the other was fixed and H&E-stained. DNA content abnormalities were defined as the presence of aneuploidy (hypo-, hyper- and tetraploidy) or an increased percent (>6%) of cells in the G2/M phase of the cell cycle. The patients were divided into a discovery and test cohorts based on the scanning site of their biopsy slides. A Hamamatsu NanoZoomer was used at the MD Anderson Cancer Center (MDACC) site to digitalize 422 H&E slides from 127 patients at 20x (0.4548 μm/pixel), and was used as the discovery cohort, (Figure 1B). 355 Slides from 64 patients were scanned on a Leica Aperio AT2 slide scanner at 20x (0.5013 μm/pixel) at Northwestern University (NU) served as a test set to image analysis algorithms’ generalizability (Figure 1B). Incident esophageal adenocarcinoma was detected in 53 of these patients during surveillance. Histological slides with significant out-of-focus issues, air bubbles, and tissues lacking BE regions were excluded from the study. All participants provided written informed consent (Fred Hutchinson Cancer Center IRB Committee D, reg. ID 5619).

**Figure 1.**
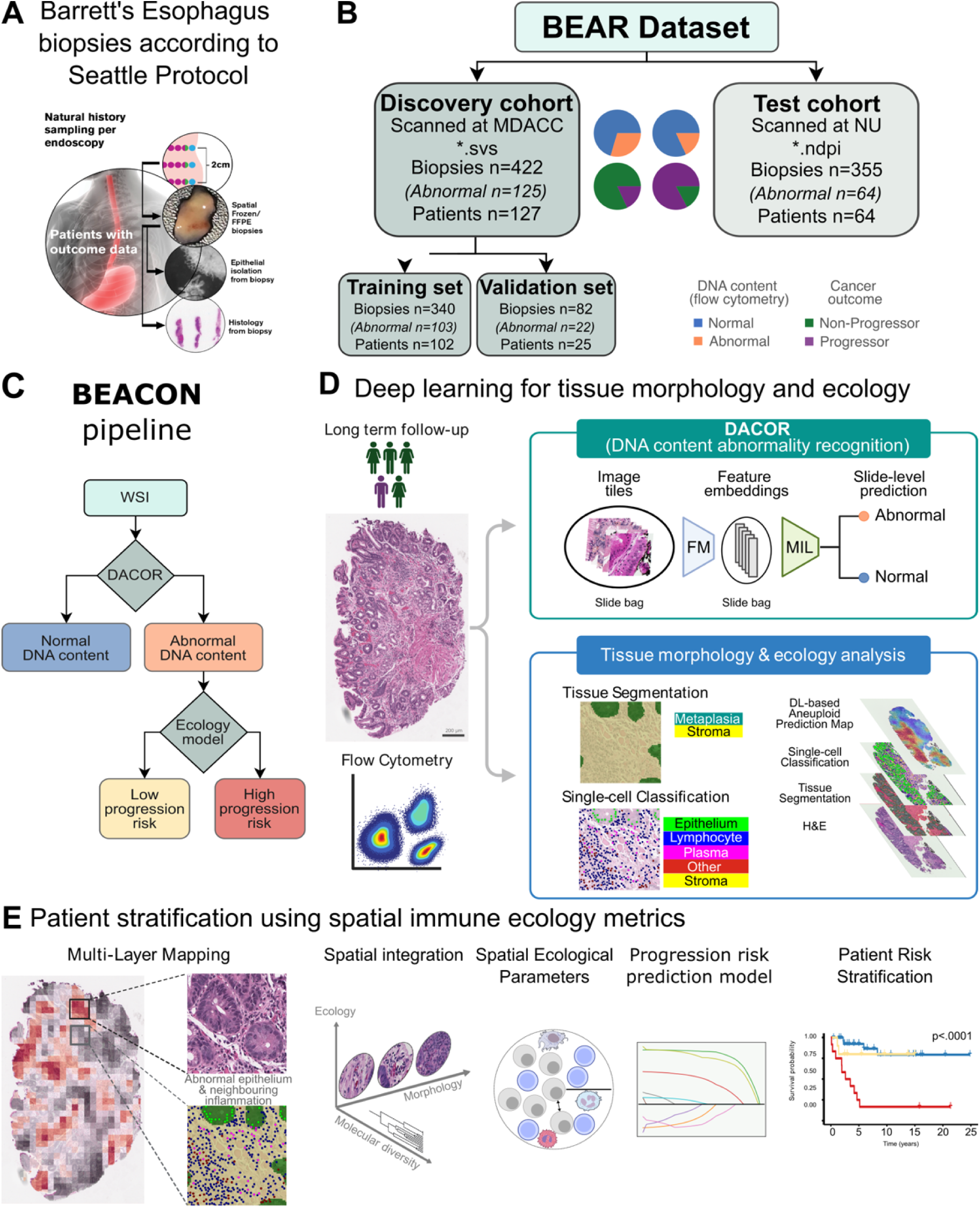
Study Overview. **A.** Systematic sampling of endoscopic biopsies, along with follow-up data and flow cytometric status, were collected to analyze the spatial ecology of BE. B. BEAR dataset was used for training an testing the models. Slides were scanned at two different sites: MDACC, NU. **C.** BEACON workflow for Barrett’s esophagus risk stratification: H&E slides are analyzed using DL models to predict DNA content abnormalities (DACOR) and characterize spatial immune ecology, enabling patient stratification by cancer progression risk. **D.** D models were trained to perform flow cytometric abnormality prediction, single-cell classification, and semantic segmentation, enabling spatial analysis of molecular and immune landscapes. Outputs from these models were integrated to generate comprehensive spatial maps. E. Ecological metrics were computed from the combined spatial data and used to stratify patients by their risk of progression to esophageal adenocarcinoma.

### DNA content prediction from tissue space

DACOR employed weakly-supervised learning using the CLAM framework.^14^

The REMEDIS (path-152×2-remedis-m) foundation model,^21^ pretrained on pathology images, as the base feature extractor. The model was fine-tuned using self-supervised learning SimCLR framework^22^ to capture robust morphological features. A dynamic region sampling strategy ensured consistent training data: for each epoch, a 1024×1024-pixel region was randomly selected from each WSI, and two non-overlapping 224×224-pixel tiles were extracted and augmented. (Supplementary Table 1) Regions and tiles were generated to contain sufficient tissue coverage (below 50% and 70%, respectively). The contrastive loss function optimized the model by maximizing cosine similarity^23^ between feature vectors of augmented tiles from the same region (positive pairs) while minimizing similarity between vectors from different regions (negative pairs). After self-supervised fine-tuning, the model extracted features from tile images.

For DACOR training, samples scanned at MDACC served as the discovery cohort, while those scanned at NU formed the test cohort. Discovery cohort samples were split into training and validation sets (85:15 ratio) in a patient-constrained manner. Architecture and training parameters are in Supplementary Tables 2-3.

Local attention scores for each slide were extracted to determine regional abnormality probabilities. Local predictions were generated using a sliding window approach with 50% overlap. This process yielded prediction scores at 112×112-pixel resolution, enabling detailed spatial mapping of DNA content abnormality probability across tissue regions.

### Complementary model trainings for microenvironment analysis

A cell nucleus instance segmentation model was trained using CellVit-plus-plus^24^ on 40 training, 10 validation, and 20 test slides randomly selected from the cohorts. Ground truth annotations to detect cell nucleus borders and distinguishing epithelial cells from other cells were created by one pathologist (C.E.) in QuPath. (Supplementary Table 4) The pretrained Virchow foundation model^25^ was fine-tuned (Supplementary Tables 5-6), and model performance was evaluated using intersection over union with >50% overlap threshold. QuPath was used to extract nuclear features including intensity (n=5), Haralick texture^26^ (n=13), and shape features (n=6) (Supplementary Table 7) for comparing DNA content normal versus abnormal epithelial cells.

Three pathologists (C.E., M.D.S., F.G.A.) annotated cells into five classes: lymphocytes, plasma cells, other immune cells (macrophages, neutrophils, eosinophils), epithelial cells, and stromal cells (fibroblasts, endothelial cells). (Supplementary Table 8) The same slide splits were used for training. The fine-tuned model from nucleus segmentation provided feature embeddings for cell classification. Training parameters are in Supplementary Table 9.

For tissue segmentation, a SegFormer^27^ model with MiT-B0 architecture was trained to distinguish columnar epithelium, stroma, squamous epithelium, and background. The model was pre-trained on annotated TCGA WSIs^28^ (1,985 training tiles from 120 WSIs, 585 validation tiles from 31 WSIs), then fine-tuned on in-house biopsy annotations by one pathologist (C.E.). Data augmentation^23^ included brightness, contrast, saturation, hue, noise, and blur adjustments (details in Supplementary Table 10). The model used cross-entropy loss, Adam optimizer (learning rate 1e-4, batch size 8), and early stopping with 10-epoch patience.

### Spatial ecological computation

Predictions from DACOR, cell classification, and tissue segmentation models were integrated for spatial immune ecology analysis. Positive regions were extended to include neighboring tiles to capture peritumoral inflammation. Immune ecological metrics were calculated in R: density, Morisita-Horn index, Getis-Ord-Gi*, Ripley’s H-function, distance-based G-function, k-nearest neighbor, and Moran’s I-function. (details in Supplementary Table 11) A LASSO logistic regression model^29^ using L1 penalty (α=1.0) was trained for risk stratification on slides predicted DNA content abnormal by DACOR. Cross-validation selected the optimal lambda from 100 logarithmically spaced values (0.001-1.0) using the 1 standard error rule. Patient-level risk scores were determined by maximum slide score, with optimal threshold from receiver operating characteristic curves on the training set. To determine the optimal threshold for risk stratification, all the slides were scored by the model for their risk and the scores aggregated for the max score at the patient level.

### Survival analysis and other statistical methods

Student’s t-tests compared age, sex, flow cytometry status, and progression; Pearson’s chi-squared tests were used for other comparisons. Nuclear and immune features were compared using two-sided t-tests. Statistical significance was set at P<0.05.

All the plots were generated using the ‘ggplot2’ packages and statistical significance was calculated using ‘ggpubr’ package’. Survival tests were conducted using Kaplan–Meier estimator (‘ggsurvplot’ R function from the ‘survminer’ and ‘survival’ R packages) as well as Cox model (‘coxph’ R function and displayed using ‘ggforest’ R function). To account for multiple testing in the multivariate Cox regression analysis, we applied Bonferroni correction by dividing the significance threshold (α = 0.05) by the number of features tested in the ecology metrics significance analysis. Clinical parameters in multivariate models included age, sex, BE length, and dysplasia status. Patients with concurrent malignancies (22 test cohort, 2 discovery cohort) were excluded from survival analyses. All analyses were conducted in R v.4.3.3.

## Results

### Study overview

In this study, we leveraged DL techniques to discover molecular and immune interactions relevant to BE patient prognosis and to determine cancer progression risk, using routine pathology endoscopic biopsy slide images. Our study utilized a subset of the unique BEAR dataset (Figure 1A, Table 1), comprising 777 biopsies from 191 BE patients. The cohort was split into a discovery and a test cohort, based on the site of digitalization (the MDACC and the NU, Figure 1B).

**Table 1.** Demographics, Clinical, and Molecular Characteristics of Study Population.

| Variable | Biopsy-level |  |  | Patient-level |  |  |
| --- | --- | --- | --- | --- | --- | --- |
|  | Discovery | Test | *p*-value | Discovery | Test | *p*-value |
| Age, mean (SD) | 63.9 (10.5) | 69.6 (8.8) | 2.05e-15 | 62.1 (10.9) | 70.9 (9.1) | 2.88e-08 |
| Sex, n (%) |  |  | 1.32e-16 |  |  | 4.45e-02 |
| Female | 124 (29.4%) | 21 (5.9%) |  | 31 (24.4%) | 7 (10.9%) |  |
| Male | 298 (70.6%) | 334 (94.1%) |  | 96 (75.6%) | 57 (89.1%) |  |
| Flow cytometry, n (%) |  |  | 2.45e-04 |  |  | 8.99e-01 |
| Normal | 297 (70.4%) | 291 (82%) |  | 86 (67.7%) | 42 (65.6%) |  |
| Abnormal | 125 (29.6%) | 64 (18%) |  | 41 (32.3%) | 22 (34.4%) |  |
| Progression, n (%) |  |  | 1.66e-58 |  |  | 2.52e-17 |
| NCO | 302 (86.5%) | 48 (13.5%) |  | 104 (81.9%) | 11 (17.2%) |  |
| CO | 120 (28.4%) | 307 (86.5%) |  | 23 (17.6%) | 53 (82.8%) |  |
| BE length, mean (SD) | 7.4 (4.1) | - |  | 5.8 (3.9) | - |  |
| Follow-up month, mean (SD) | 98.1 (76.6) | 25.8 (70.8) | 3.74e-38 | 113.9 (77.8) | 23.9 (83.5) | 6.46e-11 |
| Biopsy per patient, mean (SD) |  |  |  | 3.3 (2.7) | 5.5 (3.5) | 2.20e-05 |

NCO: Non-cancer outcome, CO: Cancer outcome, SD: Standard deviation. *p*-value was calculated for sex, flow cytometry and progression with student's *t*-test, for the rest we used Pearson's chi-squared test.

Our BEACON (Figure 1C) approach comprises three integrated key steps: (1) training DACOR to predict DNA content abnormalities from H&E slides using flow cytometry as ground truth (Figure 1D); (2) developing single-cell classification and tissue segmentation models to characterize spatial interactions between DNA content abnormal cells and the tissue microenvironment, integrating these with DACOR’s localized predictions; and (3) computing immune ecological metrics from this spatial information and trained a risk stratification model to classify patients by progression risk (Figure 1E).

### Multi-instance learning for DNA content predictions from histology images

DACOR processes H&E-stained slide images through two main steps: feature embedding generation from image tiles and feature aggregation for slide-level predictions.

To achieve robust feature extraction, we employed the REMEDIS foundation model, which was pretrained on pathology images. The foundation model underwent additional self-supervised training to ensure context-relevant feature extraction from H&E images (Figure 2A). WSIs were tiled into image patches, and feature embeddings were generated for each tile using the trained encoder. These tile embeddings were collected into slide-level bags that served as input for MIL-based DACOR model, training.

**Figure 2.**
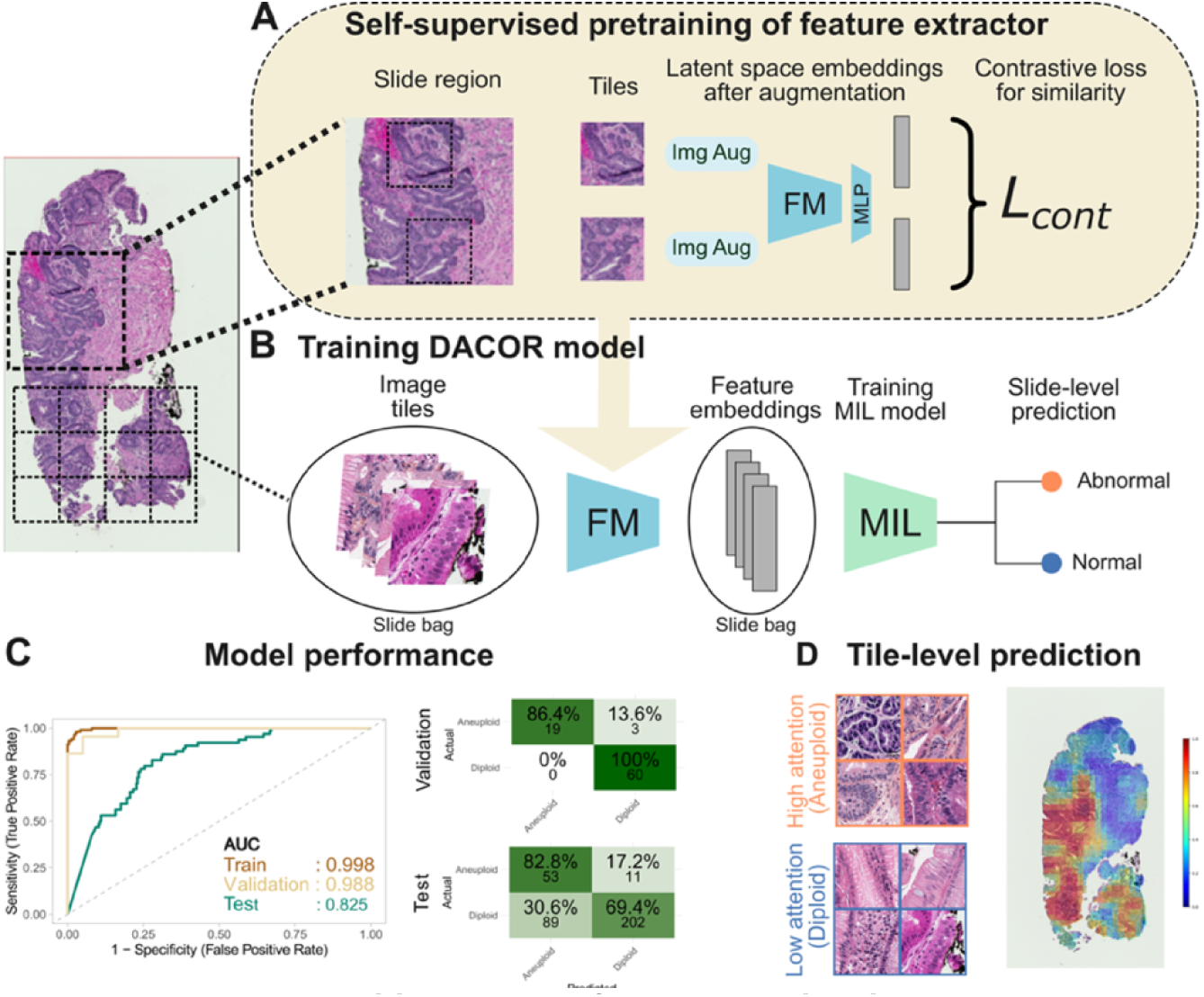
DACOR, DNA content recognition, model from H&E slide images. **A.** Self-supervised fine-tunin workflow using the SimCLR framework to adapt the REMEDIS foundation model for context-relevant featur extraction from H&E-stained tissue images. **B.** The DACOR model for DNA content abnormality works directly from pathology whole-slide images, demonstrating strong predictive performance through attention-based featur aggregation. **C.** The model reached a balanced accuracy of 93.2% (validation dataset) and 76.1% (test dataset), with corresponding AUC values of 0.988 and 0.825, respectively. The primary cause of reduced test performance wa attributed to false positives. **D.** Qualitative analysis of high-attention versus low-attention tissue regions, revealing morphological differences between DNA content abnormal and normal areas, including increased nuclear pleomorphism, hyperchromasia, and irregular crypt architecture in DNA content abnormal regions. Img Aug: Image augmentation, FM: Foundation model, MLP: Multilayer perceptron, L_cont_: Contrastive loss, MIL: Multi-instance learning, AUC: Area under curve, AUC: Area under curve,

The DACOR model was trained to predict DNA content abnormalities in BE based using flow cytometry results as ground truth for slide-level labels (Figure 2B). The discovery cohort was split into training and validation subsets at an 85:15 ratio. Images from the training set images were used to train the model, while the held-out validation set images were used to monitor training performance and validate the performance of the model in the discovery cohort. The trained model demonstrated strong performance across both validation and test datasets, achieving balanced accuracies of 93.2% and 76.1% with AUCs of 0.988 and 0.825, respectively (Figure 2C).

Beyond slide-level predictions, we aimed to improve explainability and generate a better understanding of the morphological alterations that reflect DNA content abnormalities. We utilized the tile-level attention heatmaps of the DACOR model to identify the regions with DNA content abnormality for each biopsy, enabling investigation of associated morphological changes. Visual evaluation revealed that regions with the highest DNA content abnormality prediction scores exhibited hyperchromatic nuclei and increased mitotic activity, consistent with known manifestations of aneuploidy (Figure 2D).^30,31^

### Nuclei with DNA content abnormalities exhibit distinct morphology

We aimed to quantify DNA content abnormality-related changes in epithelial nuclei using image analysis. We trained a nuclear instance segmentation model (Figure 3A). Manual annotations from a pathologist on 70 randomly selected WSIs were used for training and testing. In addition to detecting all cells, the model was trained to select epithelial cells for further feature analysis. The model achieved dice scores of 0.830 and 0.835, and F1-scores of 0.801 and 0.763 on validation and test datasets, respectively (Figure 3B).

**Figure 3.**
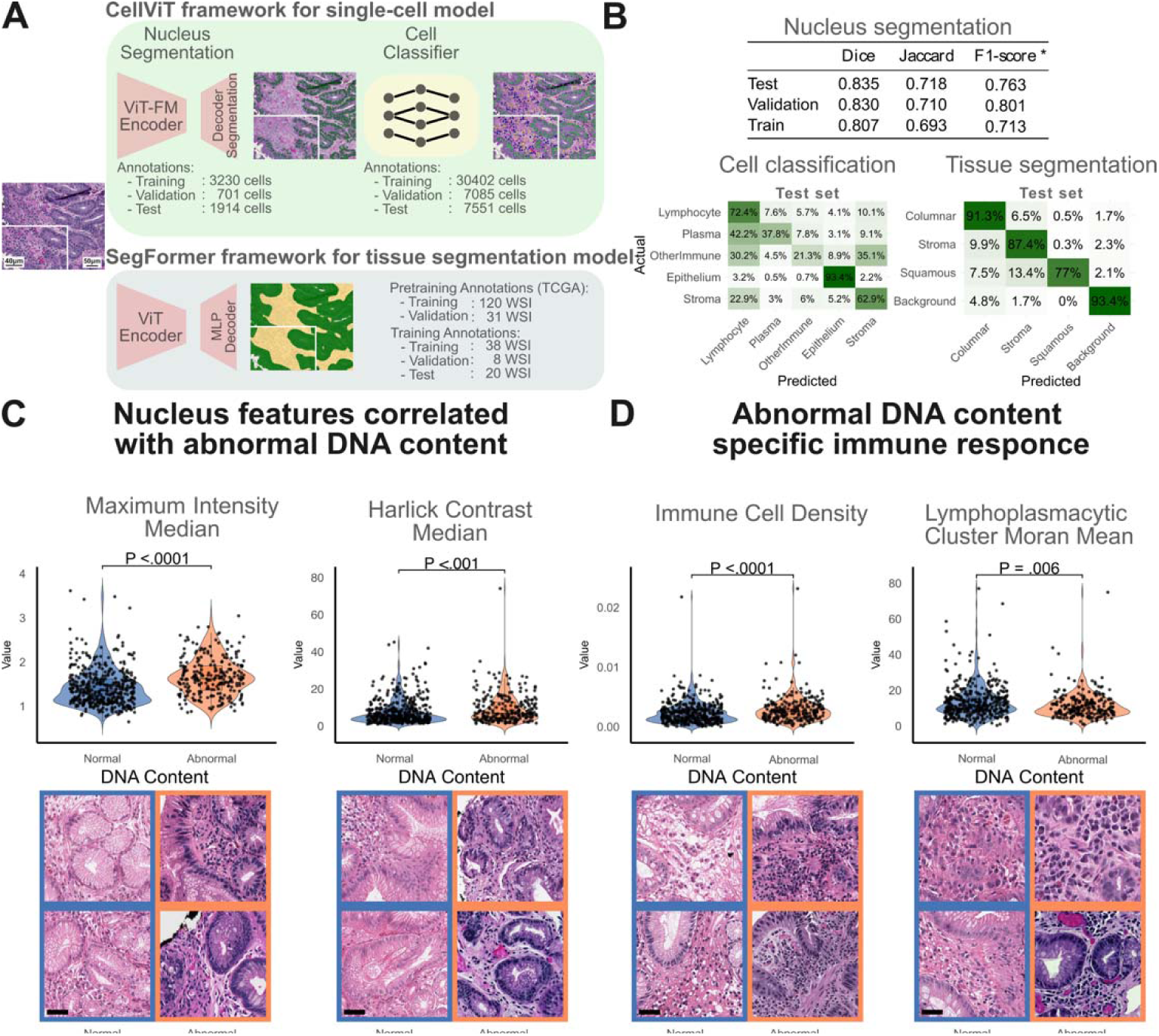
Immune landscape characterization using image analysis models. **A.** Two complementary DL models were developed to quantify the spatial immune landscape in BE biopsies. (Top) Single-cell analysis pipelin consisting of nucleus instance segmentation followed by cell classification into five distinct categories: lymphocytes, plasma cells, other immune cells, epithelial cells, and stromal cells. (Bottom) Semantic segmentation model trained to identify and segment tissue components within BE images. **B.** Performance metrics of the cell classification and tissue segmentation models. **C.** Morphological analysis of epithelial nuclei revealed significant differences between DNA content abnormal and normal regions. BE epithelial nuclei in DNA content abnormal regions demonstrate significantly higher staining intensity (left), and increased Haralick contrast, indicating enhanced nuclear pleomorphism and chromatin texture heterogeneity (right). Example images from DNA content abnormal and normal regions. (bottom) **D.** Spatial immune response analysis showing significantly increased immune cell density in DNA content abnormal regions compared to diploid areas (left). DNA content abnormal regions exhibited elevate concentrations of lymphoplasmacytic cell clusters, as quantified by Moran’s I spatial autocorrelation analysis, indicating enhanced local immune activation and inflammatory response in these areas with DNA content abnormalities (right). Example images from DNA content abnormal and normal regions. (bottom) VIT-FM: Visual transformer foundation model, scale bars: 20 μm.

Using the cell nuclei model’s outputs, we quantified morphometric changes, including size, shape, staining intensity, along with texture features in BE epithelium. To investigate DNA content abnormality-related changes, we compared features from epithelial cells between slides with abnormal and normal DNA content. Our analysis revealed that cell nucleus color intensity and nucleus size were significantly increased, while circularity was significantly decreased in DNA content abnormal cells (Figure 3C left, Supplementary Figure 1). Furthermore, the Haralick contrast feature, which represents nuclear heterogeneity and pleomorphism, and nucleus color standard deviation were significantly elevated in DNA content abnormal samples, Figure 3, Supplementary Figure 1). These findings demonstrate that DNA content abnormality-related morphological changes in epithelial cells can be quantitatively measured and characterized.

We also investigated whether DNA content abnormality-related changes could be observed within individual slides. To address this, we compared cell features between epithelial regions that contributed most and least to the model’s DNA content abnormality predictions using the model’s heatmap output. High-attention score regions (DNA content abnormality-predicted regions) showed significantly higher nucleus color intensity, increased nucleus size, decreased circularity, elevated Haralick contrast, and increased nucleus color standard deviation compared to low-attention regions in paired analysis (Supplementary Figure 1B). These intra-slide differences mirrored the changes observed between DNA content abnormal and normal samples, demonstrating consistency across different analytical approaches. These findings not only help explain the predictive factors underlying the DNA content abnormality prediction model’s performance but also support using the model’s attention-based output to identify and investigate DNA content abnormal regions within heterogeneous tissue samples.

### Regions with DNA content abnormality are associated with higher local immune response

To analyze the spatial heterogeneity of immune response in the BE and its association with DNA content abnormality, we trained two models to detect, classify, and localize immune, epithelial, and stromal cells within biopsies. We trained a single-cell model for cell detection and classification, and a segmentation model for determining tissue component regions. Combining predictions from these models with the local DNA content abnormality probabilities enabled precise mapping of immune response in the DNA content abnormal landscape.

This single cell model detects and classifies cells into five categories: epithelium, lymphocyte, plasma cell, other immune cells (neutrophils, eosinophils, macrophages), and stromal cells. A total of 37,487 cells were annotated from the discovery set and 7551 cells from the test set. The breakdown of annotated cell classes can be found in Supplementary Table 8. The model achieved accuracies of 76.4% and 73.3% (Figure 3B).

Identifying tissue component areas is important for cell density-based spatial calculations. Therefore, we trained a semantic segmentation model to segment images into four classes: columnar epithelium, squamous epithelium, stroma, and background. The model achieved overall mean IoU performance of 0.814 and 0.813 on validation and test datasets, respectively (Figure 3B).

By combining local DNA content abnormality prediction with single-cell classification and tissue component segmentation results, we were able to analyze immune response alterations associated with DNA content abnormality and cancer progression. In the initial analysis, we examined changes in immune cell density in relation to DNA content abnormality. The analysis revealed that immune cell density was significantly increased in biopsies with DNA content abnormal compared with diploid tissue (Figure 3D left). Moran’s local I clustering score of plasma cells was significantly higher in DNA content abnormal samples than normal ones. (Figure 3D right) Furthermore immune cell hotspot presence in high-attention regions (Getis-Ord analysis) is higher in DNA content abnormal samples than diploid samples, and epithelial–immune cell attraction score is higher in DNA content abnormal samples than normal samples (Supplementary Figure 1C). This same pattern was observed within individual DNA content abnormal biopsies when comparing regions with DNA content abnormal and normal tissue (Supplementary Figure 1D). In summary, our image analysis models effectively classified cells and segmented tissue components, enabling detailed spatial analysis of immune response alterations associated with DNA content abnormality and cancer progression.

### DL-based DNA content abnormality prediction is associated with risk of cancer progression

Since DNA content abnormalities correlate with cancer progression, we investigated this association using both flow cytometry-based results and DACOR’s predicted. To enable patient-level analysis, we determined DNA content abnormality status for each patient using identical criteria across both flow cytometry results and DACOR’s predictions: a patient was classified as “DNA content abnormal” if any of their biopsies tested positive; patients with all negative biopsies were classified as having normal DNA content. This approach allowed us to assess whether DACOR predictions correlate with clinical outcomes similarly to flow cytometry.

Flow cytometry results and the model’s DNA content abnormality predictions showed 64.9% and 72.3% agreement (p-values 1.04e-3 and 6.34e-9, respectively) with cancer progression across the entire dataset (Figure 4A). Similarly, both flow cytometry and predicted DNA content abnormality status-based DNA content abnormality correlated significantly with patient progression in the discovery cohort (p-values 4.89e-4 and 1.09e-2, respectively) (Supplementary Figure 2A-left). However, the correlation was not statistically significant in the test cohort for either flow cytometry or DACOR predictions (p-values 1.12e-1 and 2.36e-1, respectively) (Supplementary Figure 2A-right), which can be related with smaller number of non-progressor patients in the cohort.

**Figure 4.**
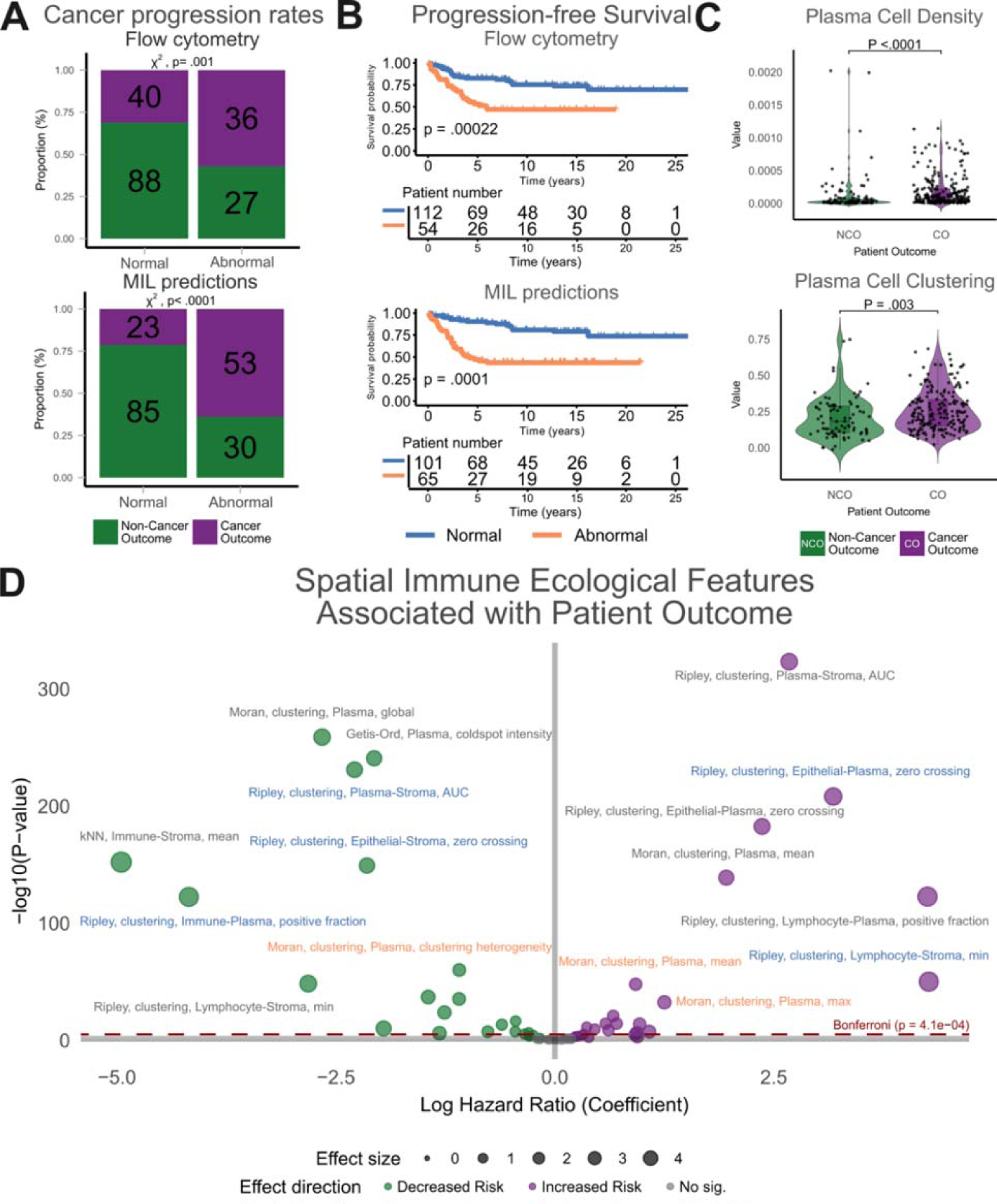
Clinical outcome analysis. **A**. Patient’s progression to carcinoma was found significantly correlated with presence of DNA content abnormality based on both flow cytometry and DACOR model’s predictions. **B.** The progression-free survival of patients was significantly correlated with DNA content abnormality based on both flow cytometry analysis and our model’s predictions. **C.** In immune ecological analysis plasma cell density and clustering based on Getis-Ord analysis was found significantly higher in cancer progressor patients within DNA content abnormal group. **D.** Volcano plot of Cox analysis of immune ecological metrics for cancer progression showed positive correlation of plasma cellular inflammation in clustering pattern around epithelial cells in DNA content abnormal, and attraction to stroma cells with cancer progression. Points above the dashed lines are metrics that are statistically significant after Bonferroni correction for multiple testing.

To further investigate the complementary function of flow cytometry and DACOR predictions in determining DNA content status, we examined correlations based on different classification criteria: patients classified as DNA content abnormal when both tests were positive versus when either test was positive. The results were similar for the discovery cohort regardless of classification criteria. However, in the test cohort, we observed significant correlation with progression only when DNA content abnormality was determined by concordance of both tests (p-value 2.82e-2) (Supplementary Figure 2B). This demonstrates that the model’s predictions are not only accurate at the slide level but can also complement flow cytometry for patient-level clinical assessment of disease progression, particularly when used in combination.

Furthermore, we investigated clinical outcome differences between patients with and without DNA content abnormality. We found significantly worse progression free survival in BE patients with DNA content abnormality by both flow cytometry and DACOR model predictions on the discovery cohort (p-values 2.2e-04, 1e-04, respectively) (Figure 4B). However, in the test cohort, while patients with DNA content abnormality showed worse prognosis than diploid patients based on both flow cytometry and DACOR predictions, the results were not statistically significant (p-values 7.2e-2, 2.1e-1, respectively) (Supplementary Figure 2C). Multivariate analysis confirmed DNA content abnormality as an adverse prognostic factor in cancer progression based on both flow cytometry and DACOR prediction analyses. (Supplementary Figure 2D).

Leveraging immune ecological metrics, we investigated prognosis-relevant immune landscape changes in samples with DNA content abnormality. Increased plasma cell inflammatory density was found to be significantly correlated with progression to cancer (Figure 4C top). Furthermore, immune cell clustering analysis using Ripley’s L and Moran’s I revealed enriched plasma cell hotspots in DNA content abnormal regions that correlated with progression to EAC (Figure 4C bottom and Supplementary Figure 3A).

To further characterize the prognostic significance of immune microenvironment features, we performed multivariate Cox regression analysis on immune ecological metrics that showed significance in univariate analysis. This analysis identified positive correlations between cancer progression and both plasma cellular inflammation clustering pattern around DNA content abnormal epithelial cells and attraction to stroma cells (Figure 4D).

In summary, our findings demonstrate a significant correlation between DNA content abnormality and increased cancer progression risk in BE patients, with both flow cytometry results and predicted DNA content abnormality status. Additionally, spatial analysis of the immune microenvironment revealed that increased plasma cell density and clustering patterns serve as prognostic indicators associated with cancer progression in DNA content abnormality regions, providing insights into the immune ecological features that accompany DNA content abnormalities in BE.

### Integration of DNA content abnormality and immune spatial ecology for risk stratification

Given the significant correlations between immune ecological parameters and patient outcomes, we developed a predictive model utilizing spatial immune and morphological metrics to stratify cancer progression risk in patients with DNA content abnormality. For this aim, we utilized comprehensive immune ecological parameters, including cell density, co-localization (Morisita-Horn index), hotspot analysis (Getis-Ord-Gi*), attraction analysis (Ripley’s L-function), neighborhood analysis (distance-based G-function, k-nearest neighbor), and clustering (Moran’s I-function) together with epithelial cell nuclear features. These parameters were calculated for DNA content abnormal and normal regions, and overall tissue areas within each slide. Additionally, cell-to-cell relationship metrics were computed for all combinations between epithelial, immune, and stromal cells (a complete list of metrics in Supplementary Table 11).

The spatial metrics served as input features to train a logistic regression model using the LASSO method for risk stratification in DNA content abnormal samples. LASSO automatically selects highl1linfluence variables and downl1lweights or drops less important ones by penalizing predictor coefficients, which reduces overfitting and yields more stable, interpretable models. Due to prognostic differences between the discovery and test cohorts, particularly the unusually high cancer progression rate (82.8%) in the test cohort (Table 1), we created new training and test splits specifically for the ecology-based risk-stratification model. These new splits combined images from both the discovery and test cohorts to ensure balanced representation. The split was performed in a patient-constrained manner, balancing DNA content anomalies and the cancer progression distributions between the cohorts (Supplementary Table 12). The model achieved 86.7% and 81.7% balanced accuracy and 0.902, 0.817 AUC on training and validation subsets, respectively (Figure 5A).

**Figure 5.**
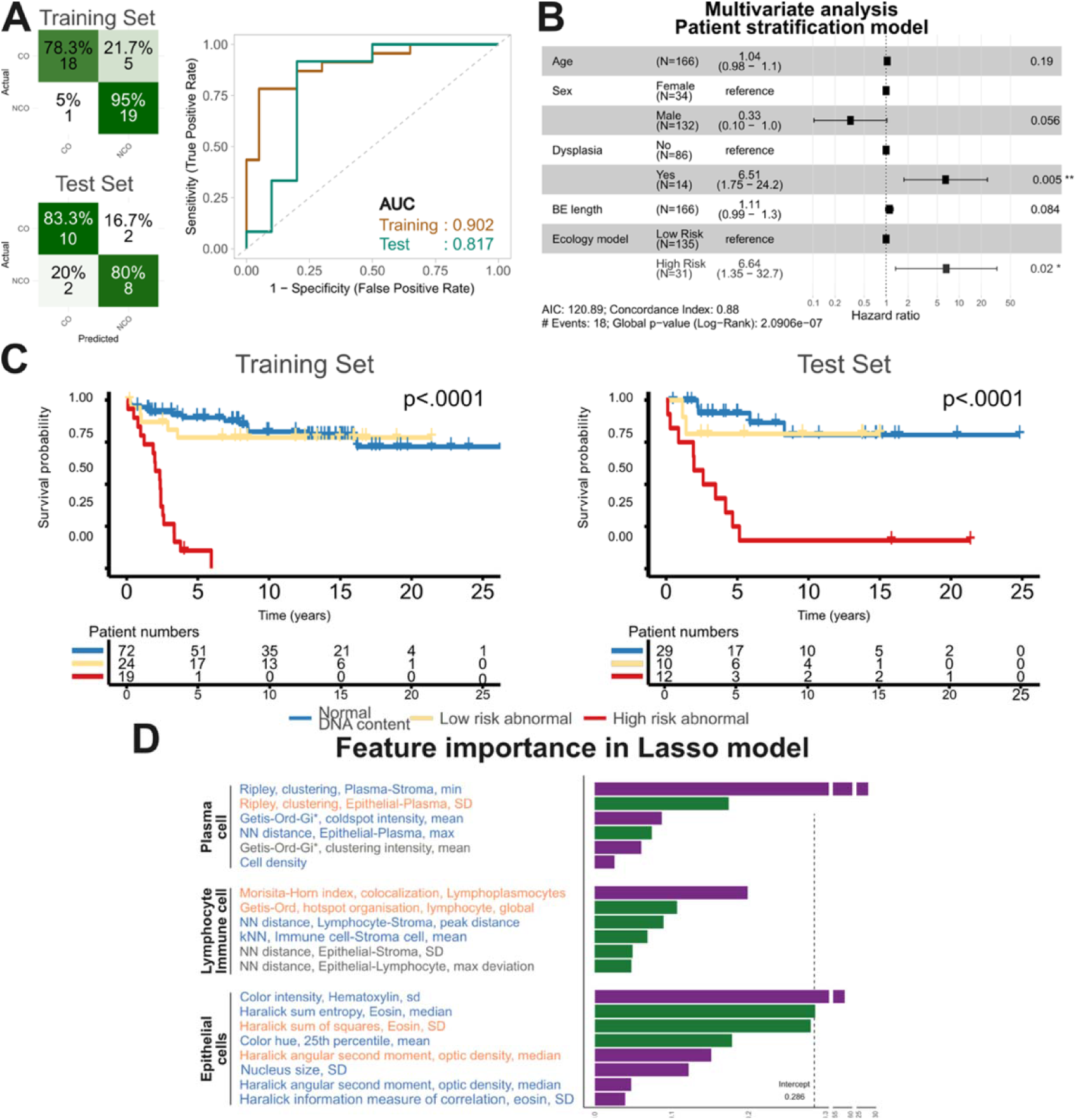
Ecological metrics predict cancer progression risk in BE with DNA content abnormality. **A.** A logistic regression model trained on DNA content abnormal samples achieved moderate classification performance for predicting cancer progression risk. The patient stratification model reached a balanced accuracy of 78.7% on th training set and 71.4% on the test set. B. The combination of DACOR model and ecology risk model yields significantly higher progression risk compared to individual models. **C.** Stratifying patients based on the presence of DNA content abnormality and high-risk ecological features yielded significant separation in progression-free survival (p < 0.0001). **D.** Feature importance analysis of the LASSO model for patient risk stratification. NN: nearest neighbor, kNN, k-nearest neighbors, SD: Standard deviation,

Cox regression analysis showed significant correlation with the predicted risk category with cancer progression for the training (HR = 9,17, p < 0.001) and test splits (HR = 5.53, p = 0.028). (Supplementary Figure 3B). Multivariate cox regression analysis revealed significant hazard ratios for the predicted risk groups (HR = 6.64, p = 0.02) for the combined whole dataset (Figure 5B). Additionally, survival analysis showed significant difference between low risk and high-risk groups for training and test datasets (p-value 1e-4, 1.4e-2, respectively) (Supplementary Figure 3C).

Notably, DNA content abnormal samples classified as low-risk by our model showed survival rates similar to patients without predicted DNA content abnormality in both training and test sets (Figure 5C), suggesting effective risk stratification within the DNA content abnormal population. Based-on the feature importance analysis of the LASSO model, the high risk group is characterized by co-localization of lymphoplasmacytic inflammation with epithelial cells with DNA content abnormality using the Morisita-Horn index, as well as increased plasma cell clustering in stroma regions distant from DNA content abnormal areas (Figure 5D). In contrast lymphocyte hotspot organizations in DNA content abnormal regions. These findings indicate not only the localization and cellular components of the immune response but also organization pattern can be an important indicator to determine prognostic risk.

In summary, our immune ecology-based model effectively stratified patients by cancer progression risk, with strong predictive performance demonstrating significant survival differences between risk groups and revealing the potential for refined prognostic assessment in BE patients with DNA content abnormality.

## Discussion

This study demonstrates that integrating molecular predictions with immune spatial ecology can effectively stratify BE patients by cancer progression risk using routine pathology slides. The evolution of neoplasm involves both molecular alterations and immune response dynamics, which can act as a selective pressure, barrier, or promoter of malignancy.^32^ Our image-based approach, BEACON, offers objective, reproducible, and explainable risk assessment without requiring additional tissue or specialized testing.

In this study, we successfully predicted DNA content abnormality from routine H&E slides by using DACOR, with significant correlation with patient outcomes. Unlike flow cytometry, our approach preserves spatial context, enabling investigation of the intricate spatial relationships between molecular alterations in the epithelium and immune response. Previous studies utilized predefined nuclear morphological features for aneuploid prediction.^34,35^ In contrast, our approach advances this by leveraging foundation model generalizability while retaining explainability, achieving significant performance improvement through automated feature learning and selective analysis of relevant tissue regions. Through spatial probability mapping, we quantitatively validated microscopic features known to be associated, including hyperchromasia, pleomorphism, and increased nuclear size.^30,31,36^ Notably, these features align with the established criteria pathologists use to diagnose dysplasia, reflecting the close relationship between dysplasia and aneuploidy. Therefore, our method systematically and objectively quantifies the same morphological indicators that have long guided clinical diagnosis. While examining DNA content-dysplasia relationships was beyond this study’s scope, localized heatmaps could direct pathologists’ attention to regions requiring examination. While attention-based heatmaps have known limitations and alternative approaches exist,^37^ they remain widely used due to computational efficiency and relative robustness. A key limitation is the lack of validation against region-specific molecular ground truth, which would strengthen confidence in the spatial resolution of our prediction capabilities.

The interaction between molecular alterations of the epithelium and surrounding immune response is a co-evolutionary process, which underscores the importance of spatially aware analyses for deriving biological insights and predicting patient risk.^18^ Recent spatial proteomics findings^38^ showing increased plasma cell interactions across progression stages align with our observation of enhanced plasma cell attraction in progressor samples. Importantly, our spatially resolved analysis provides superior understanding compared to bulk tissue analysis, revealing that immune response heterogeneity and spatial organization are critical prognostic factors.

The effect of immune response on cancer development was found to be spatial and molecular context-dependent in different tumors.^39,40^ We found that specific plasma cell clustering patterns adjacent to DNA content-abnormal regions predict progression, building on established knowledge of plasma cells in BE’s inflammatory landscape.^41^ While cross-sectional studies consistently show increased plasma cell fractions from non-tumor epithelium from BE through EAC in paired samples,^38,42^ conflicting results exist.^16^ Lin and Hickey et al^16^ found decreased plasma cell fractions but increased cell-cell attraction patterns during progression, highlighting the complexity of immune dynamics. Our spatial ecological analysis reconciles these findings by demonstrating that organization patterns, not just density, determine prognostic significance.

DL-based risk stratification in gastrointestinal research has demonstrated strong predictive performance but can suffer from limited interpretability.^43^ We developed an interpretable risk stratification model combining DNA content abnormality predictions with immune ecological metrics, achieving a hazard ratio of 6.64 (95% CI: 1.35-32.7) for progression-free survival. Unlike pan-cancer approaches using generic features,^17^ our pipeline is specifically tailored for precancerous lesion assessment, integrating disease-specific molecular alterations with detailed spatial immune analysis.

Several limitations merit consideration. First, the test cohort’s enrichment for progressors (82.8%) limits generalizability, though we mitigated this through balanced resampling for the ecology-based model. Prospective validation in independent, clinically representative cohorts will be essential to confirm model generalizability and establish clinically actionable risk thresholds. Second, using flow cytometry as ground truth introduces limitations: it was performed on one biopsy half while the adjacent half was imaged, potentially introducing errors given tissue heterogeneity. Additionally, flow cytometry requires >10% DNA content difference (∼300 Mbp) for confident abnormality determination.^33^ While our model’s clinical correlation is promising, validation with more sensitive methodologies is needed. Third, while our approach characterized nuclear morphometric features associated with DNA content abnormalities, we did not extend to broader epithelial architectural abnormalities or BE subtyping. Future work incorporating these tissue-level features alongside nuclear morphometrics, molecular predictions, and immune signatures could provide a more comprehensive characterization. Finally, establishing causality between immune phenotypes and progression requires longitudinal tracking rather than static observation. The fundamental question remains: Do immune patterns merely reflect epithelial aggressiveness, or do intrinsic immune properties determine progression trajectory? Experimental models tracking these interactions over time are needed to untangle this co-evolutionary process.

In conclusion, BEACON integrates molecular predictions with immune spatial ecology for cancer risk stratification using routine pathology. DACOR detects DNA content abnormality from H&E slides and achieves strong clinical correlation. Through spatial analysis, we identified morphological alterations and immune changes associated with progression risk. Integration of DNA content predictions with immune ecology enabled effective stratification, revealing that patients with DNA content abnormalities and high-risk immune patterns face substantially increased progression risk.

This scalable, tissue-preserving integrated computational pathology approach leverages routine specimens without additional testing for risk stratification. By combining molecular prediction with immune spatial analysis, BEACON provides objective, explainable risk assessments that could improve clinical decision-making and reduce unnecessary surveillance. By combining molecular prediction with immune spatial analysis, BEACON provides objective, explainable risk assessments that could improve clinical decision-making and reduce unnecessary surveillance. As computational pathology advances, such integrated multi-scale approaches may transform cancer risk assessment and patient management across precancerous conditions.

## Data availability

The annotations and paired H&E image tiles used for training and testing the cell nucleus instancel7lsegmentation, celll7lclassification, and tissue component segmentation models are available at zenodo upon publication of peer-reviewed paper.

## Code availability

Training and inference code for the models will be available at https://github.com/caner-ercan/BEACON upon publication of peer-reviewed paper.

## Author contributions

C.E.: Conceptualization, pathology annotation, pathology expertise, design of methodology, data processing, model development, coding, formal analysis, visualization, writing – original draft, writing – review and editing.

X.P.: Conceptualization, design of methodology, writing – review and editing.

T.G.P.: Data acquisition and curation, writing – review and editing.

M.D.S.: Pathology annotation, pathology expertise, writing – review and editing.

F.G.A.: Pathology annotation, writing – review and editing.

W.M.G.: Data acquisition and curation, writing – review and editing.

C.M.: Original idea and supervision, conceptualization, writing – review and editing.

Y.Y.: Original idea and supervision, conceptualization, design of methodology, writing – review and editing.

## Acknowledgements

This study was supported by Postdoc.Mobility fellowship (C.E.) from Swiss National Science Foundation (project number P500PM_214162). This work is supported by Spatial Ecology & QUantitative pathOlogy Image Analytical platform (SEQUOIA) through the MD Anderson STrategic Research Initiative Development Program (STRIDE). C.C.M. was supported by NIH grants 1R01CA285517-01A1 and R01CA140657. T.G.P. was supported by NIH grants U2C-CA271902, R21CA2259687 and R01CA140657. M.D.S. was supported by NCI grant R37CA269649 and DoD grant CA240129.

