## Supplementary figures and images for "Histopathology-based Spatial Profiling of Immune and Molecular Features Predicts Cancer risk in Barrett’s Esophagus"

### Supplementary Figure 1

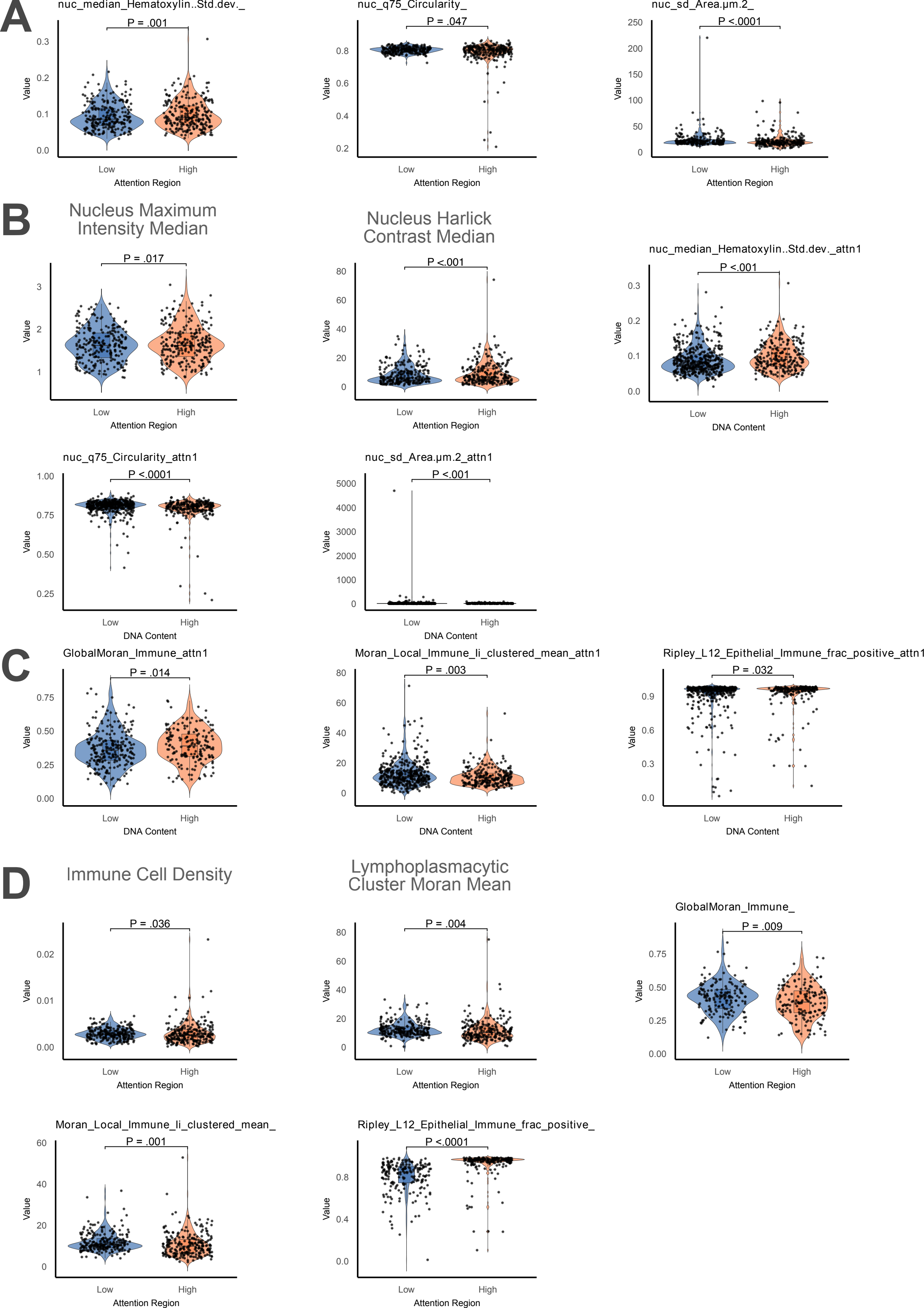

### Supplementary Figure 2

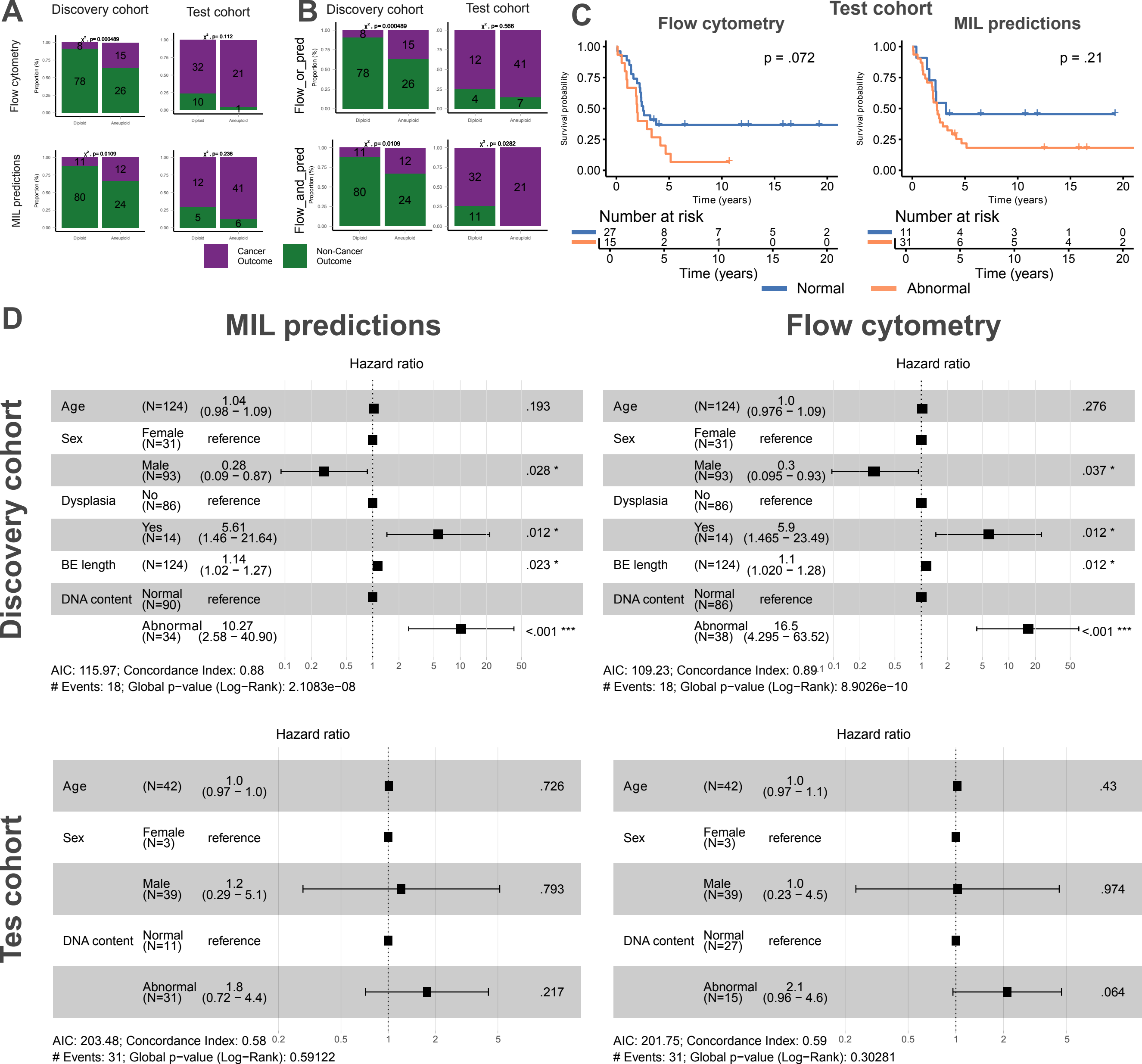

### Supplementary Figure 3

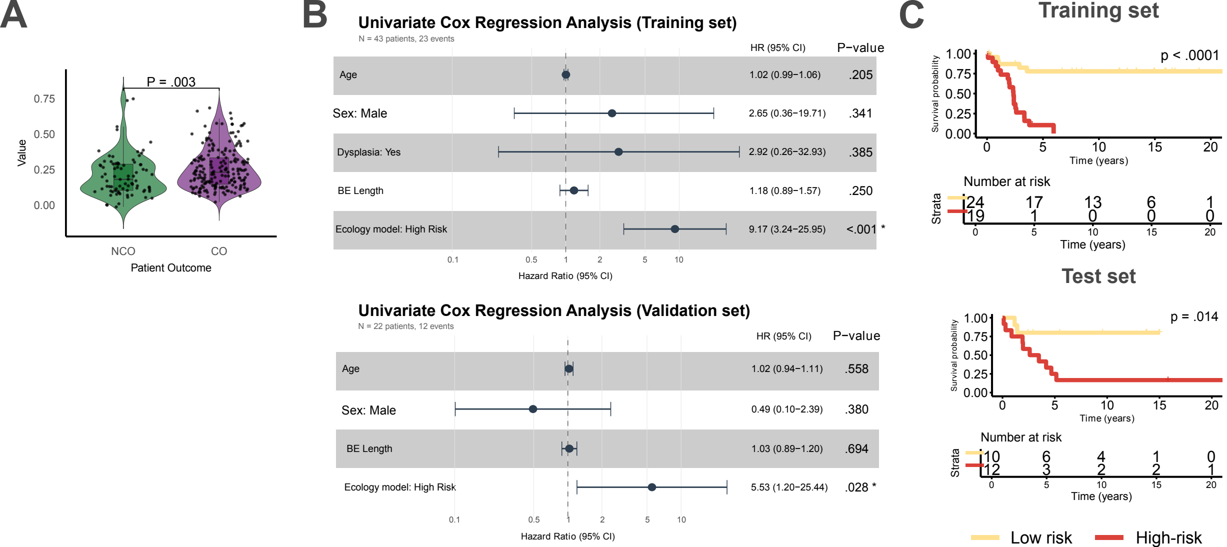
