## Supplementary Table 1 for "Histopathology-based Spatial Profiling of Immune and Molecular Features Predicts Cancer risk in Barrett’s Esophagus"

Supplementary Table 1 Simsiam transformations

| **Function** | **Scale** | **Probability** |
| --- | --- | --- |
| Horizontal Flip | - | 0.5 |
| Vertical Flip | - | 0.5 |
| Color Jitter | Brightness: 0.4 Contrass: 0.4 Saturation: 0.4 Hue: 0.1 | 0.8 |
| Gaussian Blur | limits:[3:7] | 0.7 |
| To gray |  | 0.05 |
