## Supplementary Table 2 for "Histopathology-based Spatial Profiling of Immune and Molecular Features Predicts Cancer risk in Barrett’s Esophagus"

Supplementary Table 2: MIL model architecture for DACOR

| Block | Activation | Drop-out | Input size | Output size | Notes |
| --- | --- | --- | --- | --- | --- |
| Linear block | ReLU | 0.25 | Nx1024 | Nx512 | Initial feature transformation |
| Attention network gated (a) | Tanh | 0.25 | Nx512 | Nx384 | Attention branch A |
| Attention network gated (b) | Sigmoid | 0.25 | Nx512 | Nx384 | Gating branch B |
| Attention network gated (c) | - | - | Nx384 | Nx2 | Final attention weights |
| Bag classifiers | - | - | Nx512 | 2x1 | Sample level classification |

*: N represents the number of patches for each WSI (slide bag)
