## Supplementary Table 3 for "Histopathology-based Spatial Profiling of Immune and Molecular Features Predicts Cancer risk in Barrett’s Esophagus"

Supplementary Table 3 MIL model training parameters for DACOR

| **Category** | **Parameter** | **Value** |
| --- | --- | --- |
| Loss functions | Instance loss function | Focal-cross entropy |
|  | Optimizer | Adam |
|  | Positive class weight | 1.0 |
| Bag features | Bag size | 8 |
|  | Bag loss function | Cross-entropy |
|  | Bag loss weight | 0.5 |
| Early stopping | Max epoch | 200 |
|  | Patience | 20 |
| Learning Rate schedule | Patience | 8 |
|  | Cooldown | 10 |
|  | Factor | 0.5 |
|  | Monitor | Validation loss |
|  | Initial learning rate | 1e-4 |
