## Supplementary Table 4 for "Histopathology-based Spatial Profiling of Immune and Molecular Features Predicts Cancer risk in Barrett’s Esophagus"

Supplementary Table 4 Nucleus instance segmentation finetuning annotations

| **Split** | **Epithelial cells** | **Other cells** | **Total** |
| --- | --- | --- | --- |
| Training | 3095 | 135 | 3230 |
| Validation | 657 | 44 | 701 |
| Test | 1838 | 76 | 1914 |
