## Supplementary Table 5 for "Histopathology-based Spatial Profiling of Immune and Molecular Features Predicts Cancer risk in Barrett’s Esophagus"

Supplementary Table 5 Cell nucleus instance segmentation model parameters

| Parameter | Value |
| --- | --- |
| Random seed | 19 |
| Stain normalisation | TRUE |
| Input shape | 256x256 px |
| Backbone | Virchow |
| Batch size | 8 |
| Drop rate | 0.1 |
| Optimizer | AdamW |
| Unfreeze decoder epoch | 15 |
| Unfreeze encoder epoch | 35 |
| Early stopping patience | 30 |
| Early stop monitor metric | Validation loss |
| Maximum epoch | 150 |
| Initial learning rate | 0.001521 |
| Weight decay | 0.0003624 |
| Learning rate schedullar function | Constant |
