## Supplementary Table 6 for "Histopathology-based Spatial Profiling of Immune and Molecular Features Predicts Cancer risk in Barrett’s Esophagus"

Supplementary Table 6 Random transformation parameters of CellVit models’ training

| **Function** | **Value** | **probability** |
| --- | --- | --- |
| Random rotation | 90 degree | 0.5 |
| Horizontal flip | - | 0.5 |
| Vertical flip | - | 0.5 |
| Downscale | Scale: 0.1 | 0.15 |
| Blur | Limit: 10 | 0.2 |
| Gauss noise | Limit: 50 | 0.25 |
| Color jitter | Brightness: 0.25 Contrass: 0.25 Saturation: 0.1 Hue: 0.05 | 0.2 |
| Superpixels | N_segments: 200 | 0.1 |
| Zoom blur | Min dimention: 128 | 0.1 |
