## Supplementary Table 7 for "Histopathology-based Spatial Profiling of Immune and Molecular Features Predicts Cancer risk in Barrett’s Esophagus"

Supplementary Table XXX Nuclear texture feature quantifications

| **Feature Code** | **Name** | **Description** | **Biological Interpretation** |
| --- | --- | --- | --- |
| **F0** | Angular Second Moment | texture uniformity | Chromatin homogeneity; uniform nuclei have high values |
| **F1** | Contrast | texture contrast | Chromatin intensity variation; heterochromatin vs euchromatin |
| **F2** | Correlation | texture correlation | Spatial organization of chromatin patterns |
| **F3** | Sum of Squares | texture variability | Overall chromatin intensity dispersion |
| **F4** | Inverse Difference | texture homogeneity | Local chromatin uniformity; smooth vs coarse texture |
| **F5** | Sum Average | intensity average | Mean chromatin density across texture patterns |
| **F6** | Sum Variance | intensity variance | Variability in local chromatin density |
| **F7** | Sum Entropy | texture complexity | Chromatin pattern randomness and disorder |
| **F8** | Entropy | randomness | Overall chromatin structural disorganization |
| **F9** | Difference Variance | edge variance | Variability in chromatin boundaries and transitions |
| **F10** | Difference Entropy | edge entropy | Randomness in chromatin boundary patterns |
| **F11** | Information Measure of Correlation 1 | texture organization | Structured vs random chromatin arrangement |
