## Supplementary Table 8 for "Histopathology-based Spatial Profiling of Immune and Molecular Features Predicts Cancer risk in Barrett’s Esophagus"

Supplementary Table 8 Annotation counts for cell classification model training.

| Split | Lymphocyte | Plasma cell | Other immune cell | Epithelial | Stroma | Total |
| --- | --- | --- | --- | --- | --- | --- |
| Training | 9885 | 1644 | 1054 | 13166 | 4671 | 30420 |
| Validation | 2167 | 445 | 289 | 3303 | 881 | 7085 |
| Test | 2465 | 453 | 337 | 3354 | 942 | 7551 |
