## Supplementary Table 9 for "Histopathology-based Spatial Profiling of Immune and Molecular Features Predicts Cancer risk in Barrett’s Esophagus"

Supplementary Table 9 Cell classification model training parameters

| **Parameter** | **Value** |
| --- | --- |
| Classifier model hidden dimension | 512 |
| Initial learning rate | 0.0002028932649229723 |
| Weight decay | 0.0002820281690012052 |
| Tile size | 256x256 |
| Down sampling scale | 1 |
| Class weight list: |  |
| - Lymphocyte | 0.054777876705837836 |
| - Plasma cell | 0.21165309698443244 |
| - Other immune cells | 0.579307234396314 |
| - Epithelial cells | 0.03191575159272495 |
| - Stroma cells | 0.12234604032069071 |
| Optimizer | AdamW |
| Drop rate | 0.1 |
| Classifier hidden layer dimension | 128 |
| Stain normalization values | mean: [0.5, 0.5, 0.5] std: [0.5, 0.5, 0.5] |
| early_stopping_patience | 30 |
| Early stop monitor metric | Validation loss |
| Maximum epoch | 150 |
| Loss function | Cross entropy |
