## Supplementary Table 10 for "Histopathology-based Spatial Profiling of Immune and Molecular Features Predicts Cancer risk in Barrett’s Esophagus"

Supplementary Table 9: Random transformation parameters of SegFormer model’s training

| **Function** | **Value** | **Probability** |
| --- | --- | --- |
| random brightness adjustment | range -0.2, 0.2 | 45% |
| random contrast adjustment | range -0.2, 0.2 | 45% |
| random saturation adjustment | range:-30, 30 | 45% |
| random hue adjustment | range: -20,20 | 45% |
| random gausian noise | range: -0.2,0.44 | 20% |
| random blur | range: 3,7 | 20% |
