## Supplementary Table 11 for "Histopathology-based Spatial Profiling of Immune and Molecular Features Predicts Cancer risk in Barrett’s Esophagus"

Supplementary Table XXX Ecology metrics and summarizations

| Method | Metric | Explanation |
| --- | --- | --- |
| Ripley’s L | AUC | Cumulative clustering tendency |
|  | Mean | Average clustering tendency |
|  | Max | Peak clustering intensity |
|  | Min | Minimum clustering value |
|  | Sd | Variability in clustering pattern |
|  | Fraction positive | Proportion of distances showing clustering |
|  | Zero crossing | Distance threshold between clustering and regularity |
| Density | Cell density | Spatial concentration of cells per unit area |
| Getis Ord | Mean (positive regions) | Hotspot intensity |
|  | Mean (negative regions) | Coldspot intensity |
|  | Mean (absolute) | Clustering strenght |
|  | SD | Clustering heterogeneity |
|  | Global | Overall spatial autocorrelation |
| k-Nearest Neighbor | Mean kNN distance | Average proximity between cell types |
|  | SD kNN distance | Variability in intercellular distances |
|  | Minimum kNN distance | Closest cell-type interaction |
|  | Maximum kNN distance | Furthest nearest neighbor distance |
|  | Distance coefficient of variation | Uniformity vs clustering in tissue organization |
| Moran | Mean (positive regions) | Clustered regions intensity |
|  | Mean (negative regions) | Dispersed regions intensity |
|  | SD | Spatial autocorrelation heterogeneity |
|  | Mean (absolute) | Overall clustering magnitude |
|  | Max (absolute) | Peak spatial autocorrelation |
|  | Global | Global spatial autocorrelation index |
| Morisita-Horn Index | Morisita value | Degree of spatial colocalization |
| Nearest neighbor distance (G function) | AUC (difference) | Overall deviation from random expectation |
|  | Mean deviation (≤150m) | Average clustering tendency at medium range |
|  | Maximum deviation | Peak clustering intensity |
|  | Peak distance | Distance of maximum clustering |
|  | SD | Heterogeneity in spatial pattern |
|  | First zero crossing | Transition distance to regularity/overdispersion |
