## Supplementary Table 12 for "Histopathology-based Spatial Profiling of Immune and Molecular Features Predicts Cancer risk in Barrett’s Esophagus"

Supplementary Table XXX Patient distribution in splits for ecology model training

|  | **Discovery cohort** | | **Test cohort** | |
| --- | --- | --- | --- | --- |
|  | **NCO** | **CO** | **NCO** | **CO** |
| **Training** | 71 | 14 | 8 | 23 |
| **Test** | 32 | 7 | 3 | 9 |

NCO: Non-cancer outcome, CO: Cancer outcome
